# Psychological factors underpinning vaccine willingness in Israel, Japan and Hungary

**DOI:** 10.1101/2021.05.24.21257465

**Authors:** Robin Goodwin, Menachem Ben-Ezra, Masahito Takahashi, Lan-Anh Nguyen Luu, Krisztina Borsfay, Mónika Kovács, Wai-Kai Hou, Yaira Hamama-Raz, Yafit Levin

## Abstract

The rapid international spread of the SARS-CoV-2 virus 19 led to unprecedented attempts to develop and administer an effective vaccine. However, there is considerable vaccine hesitancy in some countries. We investigated willingness to vaccinate in three nations with historically different levels of vaccine willingness and attitudes: Israel, Japan and Hungary. Employing an ecological-systems approach we analysed associations between demographic factors and health status, individual cognitions, normative pressures, trust in government, belief in COVID-19 myths and willingness to be vaccinated, using data from three nationally representative samples (Israel, N=1011 (Jan 2021); Japan, N= 997 (Feb 2021); Hungary, N=1131 (Apr 2021)). In Israel 74% indicated a willingness to vaccinate, but only 51% in Japan and 31% in Hungary. Multigroup regression analyses indicated greater vaccine willingness amongst those who perceived benefits to vaccination, anticipated regret if not vaccinated and trusted the government. Multi-group latent class analysis of ten COVID-19 (mis)beliefs identified three classes of myths, with concerns about the alteration of DNA (Israel), allergies (Hungary) and catching COVID-19 from the vaccine (Japan) specific to vaccine willingness for each culture. Intervention campaigns should focus on increasing trust and addressing culturally specific myths while emphasising the individual and social group benefits of vaccination.

---

Psychological factors underpinning vaccine willingness in Israel, Japan and Hungary As the world-wide threat posed by the SARS-CoV-2 virus continues the development and implementation of vaccines has become pivotal for reducing mortality and morbidity and limiting spread^1^. However, both the availability of vaccines, speed of vaccination and willingness to vaccinate varies substantially across cultures and is often influenced by historical and political factors. Three countries exemplify this variation. Israel was the first country to launch a mass vaccination drive. Starting on 20th December 2020, 15% of the country’s population had received at least their first (Pfizer) vaccination within two weeks, 39% within a month^2^. Comparative surveys (late January 2021) found 73% of Israelis willing to accept a vaccine^3^. In Japan, problems with the combined diphtheria, tetanus, and pertussis and MMR vaccines during the 1970s and 1980s, and wide-spread concern about HPV vaccination in 2013, led to a risk-averse approach towards a COVID-19 vaccine^4,5^, with Japanese regulators not approving a vaccine until mid-February 2021^2^. Only 45% of the Japanese surveyed in late January 2021 indicated willingness to take an approved COVID-19 vaccine^3^. Finally, in Hungary, political controversy around early deployment of the Russian Sputnik-V and Chinese BBIBP-CorV (Sinopharm) vaccines^6-10^ contributed to a slow initial uptake of a COVID-19 vaccine^2^. Early surveys suggested only 38%^11^-45%^12^ were willing to vaccinate during the autumn of 2020, although later studies suggested increased willingness, with 72% indicating either willingness to vaccinate or having been vaccinated by March 2021^12^.

Alongside such historical-cultural factors vaccination willingness also varies by both demographic and individual, psychological differences within countries. In Israel, some religious communities were more likely to reject the vaccine^13-15^, with uptake in Israel also lower amongst women and those who considered themselves at low risk from COVID-19^16^. Vaccine willingness was also greater in Japan amongst men, as well as older Japanese and those with chronic disease risk factors^17^. Older respondents in Hungary, as well as the more educated, were more willing to vaccinate^12^. Alongside this, a large number of individual socio-psychological variables influence vaccine uptake^18-20^, although a paucity of theory-driven approaches to vaccination means there are a limited number of systematic frameworks available^21^. We draw on the three most widely employed psychological models of vaccine willingness – the Health Belief Model, the Theory of Planned Behaviour and Protection Motivation Theory^18. 22^– to suggest an ecological systems model, organised into three nested categories. This model aims to capture micro, meso and macro-system influences on vaccine willingness^19, 23, 24^[Figure 1]. The model includes (1) individual cognitions involved in decision-making (perceptions of susceptibility and severity of illness, perceived benefits or barriers to vaccination, and anticipated regret if not vaccinated^17-19, 25-27^ (2) local group influences and norms (the influence with important others, including family and friends^18, 28^) and (3) wider macro cultural factors, including communication, trust in government and health authorities^19, 23^ as well as beliefs about COVID-19 and the vaccine^11, 20, 29, 30^. We assess this ecological model during the very different vaccination drives in Israel, Japan and Hungary. Vaccine willingness is anticipated to be positively related to both threat appraisal and the ability to confront this; specifically, perceived susceptibility of contracting COVID-19, severity of COVID-19, benefits of vaccination, and anticipated regret if not vaccinated^17-19, 25, 26, 28^. Normative pressures to vaccinate are also anticipated to encourage willingness to vaccinate, while low trust, and a willingness to accept misbeliefs about vaccination, are expected to be negatively associated with willingness to vaccinate^18^. We control in these analyses for the key demographic variables of age (positively associated with vaccine willingness^18-20, 26^), education, employment (positively associated with uptake^19^) and sex (with men more willing to vaccine^17^, or less likely to express hesitancy^20^). In Israel only we also include religiosity, anticipated to be negatively associated with vaccine willingness^20^. We also include history of previous COVID-19 infection and underlying conditions indicating morbidity/mortality risk, with health problems associated with greater vaccine resistance^20^. Further details of measures are reported below and in the Supplementary Material.

**Fig 1:**
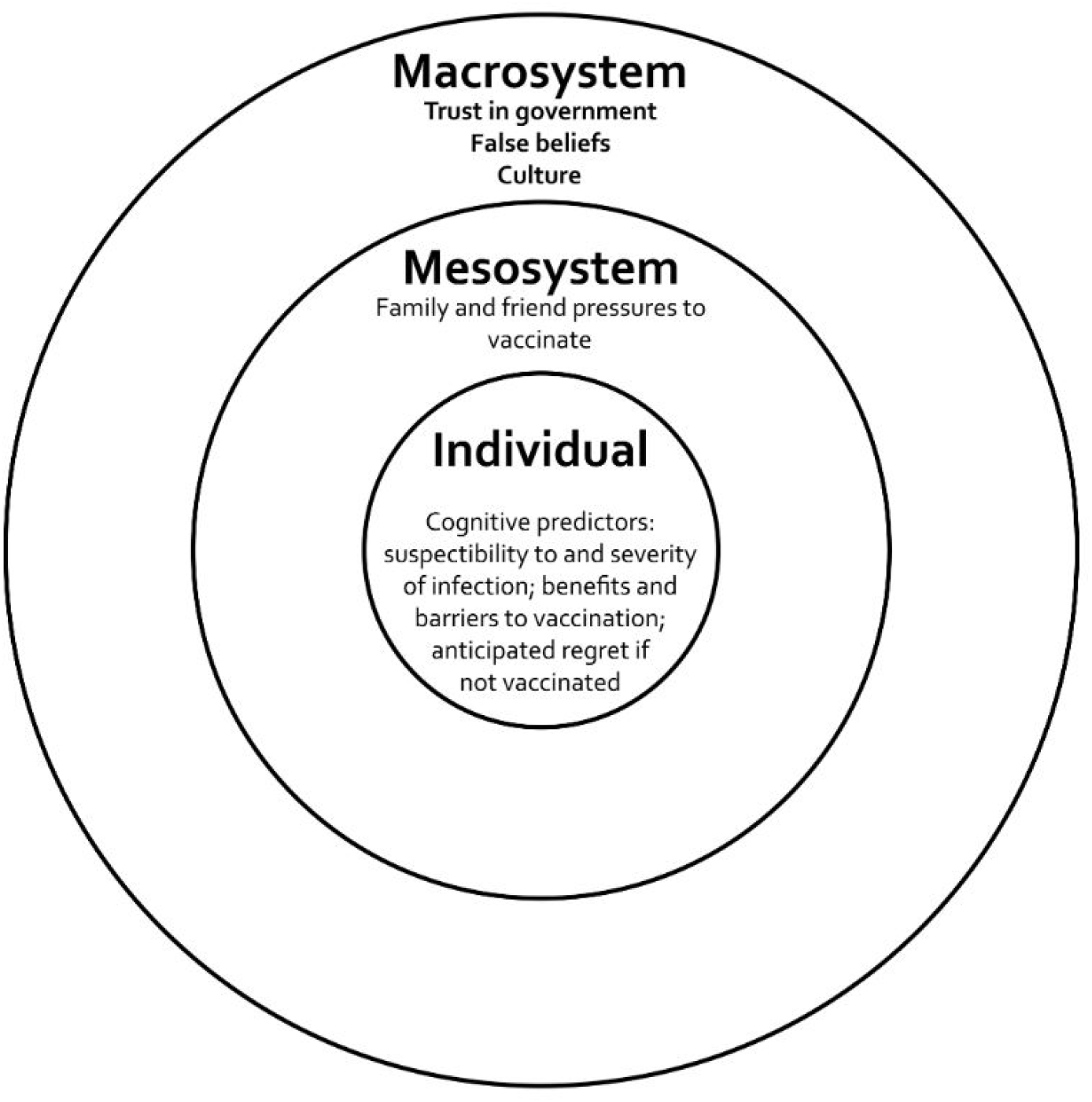
Ecological model of factors contributing to vaccine willingness

Trust in government authorities and conspiracy or false beliefs about the pandemic are often associated^31-33^. Misbeliefs, which often focus on the ‘response cost’ of vaccinations^34, 35^ are particularly important given the novelty of the vaccines developed^33, 36^, with many conspiracy theories and false beliefs about harmful effects of vaccination amplified via mass media^18, 26, 37, 38^. In US surveys (April 2020) those who adhered to vaccine conspiracy beliefs were almost four times less likely to report willingness to vaccinate [39], with similar findings reported in the UK^31, 32^, US, Spain and Mexico^32^. Such beliefs also prospectively predicted later vaccine intentions^40^. While these misbeliefs tend to correlate significantly with each other ^31, 32^, they may also be divided along dimensions^33^. In one analysis, myths about COVID-19 were divided into two, one which viewed the pandemic as a hoax, the other that suggested the human origin of the virus, with different implications for health behaviour ^41^. Other work suggests that conspiracy beliefs have consequences for behaviours that afford personal control (e.g. wearing gloves) but not for acts that threaten autonomy (which may include vaccination)^31^, while others suggest that unlike regular conspiracy accounts paranoia-like beliefs are associated with adherence to safety guidelines^42^. In our analyses we focus on common vaccine-related fears propagated across societies (e.g., about the association between vaccination and autism^43^), drawing on ten common myths identified by health advisory bodies (CDC, NHS, WHO) at the start of our first data collection. We place particular emphasis on vaccine side-effects, the most prominent concern in January 2021^44^. We test the structuring of these myths in each country via multi-group latent class analysis (LCA), and associate each with willingness to vaccinate.

This paper seeks to address four objectives. First, we compare rates of vaccine willingness in national surveys conducted in Israel, Japan and Hungary (**objective 1**). Second, we seek to understand the demographic and health risk factors associated with this willingness by considering associations between willingness and age, sex, education, employment and health status (**objective 2**). Third, we examine relative weight of each of the variables in our three-layer ecological model by conducting a multigroup path analysis in each culture (**objective 3**). Finally, (**objective 4**), we conduct a sensitivity analysis examining specific cluster structure of these ten different myths, and the impact of these on willingness to vaccinate in each country.

## Results

### Objective 1: Vaccine willingness in Israel, Japan and Hungary

Table 1 presents differences in the distribution of background and study variables between the three samples. Willingness to vaccinate was higher in Israel (74.1%) than in Japan (51.1%), or Hungary (31%) (χ^2^ (2) =397.86, P=.001) (Figure 2)

**Table 1:** Sociodemographic characteristics and variables assessed of the Israeli, Japanese and Hungarian samples (N=1011, Israel; N=997, Japan; N=1131, Hungary)

| Variable and [scale reliabilities, *r*.] (Israel, Japan, Hungary) | Frequency N (%) |  |  | Mean (SD) |  |  | Statistic test | p |
| --- | --- | --- | --- | --- | --- | --- | --- | --- |
|  | Israel | Japan | Hungary | Israel | Japan | Hungary |  |  |
| Accept the vaccine |  |  |  |  |  |  |  |  |
| - Disagree | 261 (25.8) | 448 (48.9) | 780 (69.0) | | | | $\chi^2 (2) = 397.86^{***}$ | .000 |
| - Agree | 750 (74.1) | 469 (51.1) | 351 (31%) |  |  |  |  |  |
| Demographics and health status |  |  |  |  |  |  |  |  |
| Age |  |  |  | 39.95 (14.15) | 45.63 (14.11) | 50.49 (15.95) | F(2, 3124)=134.92 | .000 |
| Sex (female) | 514 (51) | 514 (52) | 511 (33.2) | | | | $\chi^2 (6) = 13.83$ | .032 |
| Religiosity* |  |  |  |  |  |  |  |  |
| - Secular | 526 (52) |  |  |  |  |  |  |  |
| - Non-secular |  |  |  |  |  |  |  |  |
| - - Traditional | 313 (31) |  |  |  |  |  |  |  |
| - - Religious | 142 (14) |  |  |  |  |  |  |  |
| - - Ultra religious | 30 (3) |  |  |  |  |  |  |  |
| Risk group membership | | | | | | | $\chi^2 (4) = 274.803^{***}$ | .000 |
| - - Yes | 248 (24.5) | 190 (19.1) | 469 (41.5) |  |  |  |  |  |
| - - No | 763 (75.5) | 804 (80.9) | 602 (53.2%) |  |  |  |  |  |
| - - Unsure | 0 | 0 | 60 (5.3) |  |  |  |  |  |
| Education |  |  |  |  |  |  |  |  |
| - High school | 384 (38) | 473 (48) | | | | | $\chi^2 (1) = 56.19^{***}$ | .000 |
| - Student/postgrad | 627 (62) | 506 (52) |  |  |  |  |  |  |
| Personally-diagnosed with COVID-19 (yes) | 63 (6.2) | 2 (0.2) | 124 (11.1%) | | | | $\chi^2 (1) = 110.44^{***}$ | .000 |
| Family or friend member diagnosed with COVID-19 (yes) | 377 (37.3) | 11 (1.1) | 687 (60.7%) | | | | $\chi^2 (1) = 841.70^{***}$ | .000 |
| Self-rated health (4-point scale) | | | | 3.19 (.60) | 2.30 (.78) | 2.71 (.77) | F (2,3136) = 374.92*** $\eta^2 = 0.20$ | .000 |
| **Cognitive and normative factors** |  |  |  |  |  |  |  |  |
| Likelihood of infection (3 items, 5-point scale) [.73, .68 .85] | | | | 2.69 (.89) | 2.64 (.72) | 2.79 (1.04) | F (2, 3115) = 8.09*** $\eta^2 = 0.01$ | .000 |
| Perceived severity of infection (3 items, 5-point scale) [.77, .67, .80] | | | | 3.18 (.98) | 3.44 (.73) | 3.34 (1.01) | F (2, 3115) = 19.20*** $\eta^2 = 0.01$ | .000 |
| Benefits of vaccination (3 items, 5-point scale) [.79, .74, .91] | | | | 3.72 (.99) | 3.53 (.78) | 3.39 (1.22) | F (2, 3056) = 28.71*** $\eta^2 = 0.02$ | .000 |
| Barriers to vaccination (2 items, 5-point scale [*r*=.33, .47, .21]) | | | | 3.87 (1.84) | 2.95 (.91) | 2.44 (.99) | F (2, 3056) = 320.16*** $\eta^2 = 0.17$ | .000 |
| Anticipated regret (1 |  |  |  | 4.95 | 4.41 | 4.35 | F (2, 3061) | .000 |
| item, 7-point scale) | | | | (1.97) | (1.46) | (2.14) | =30.95*** $\eta^2 = 0.02$ | |
| Subjective norms (5 items, 7-point scale) [.64, .45, .66] | | | | 4.89<br>(1.08) | 4.50<br>(.82) | 4.23<br>(1.32) | F (2, 3080) =94.84*** $\eta^2 = 0.06$ | .000 |
| **Engagement, communication** |  |  |  |  |  |  |  |  |
| Trust in government (3 items, 1-5) [.82, .87, .95] | | | | 2.74<br>(1.07) | 2.57<br>(.83) | 2.41<br>(1.28) | F (2, 3087) =24.56*** $\eta^2 = 0.02$ | .000 |
| COVID-19 myths 1(no) 2 (yes) | | | | 1.22<br>(.24) | 1.28<br>(.21) | 1.25<br>(.24) | F (2, 3127) =22.98*** $\eta^2 = 0.01$ | .000 |

\*Note: Religiosity was not assessed in Japan and Hungary

**Fig. 2.**
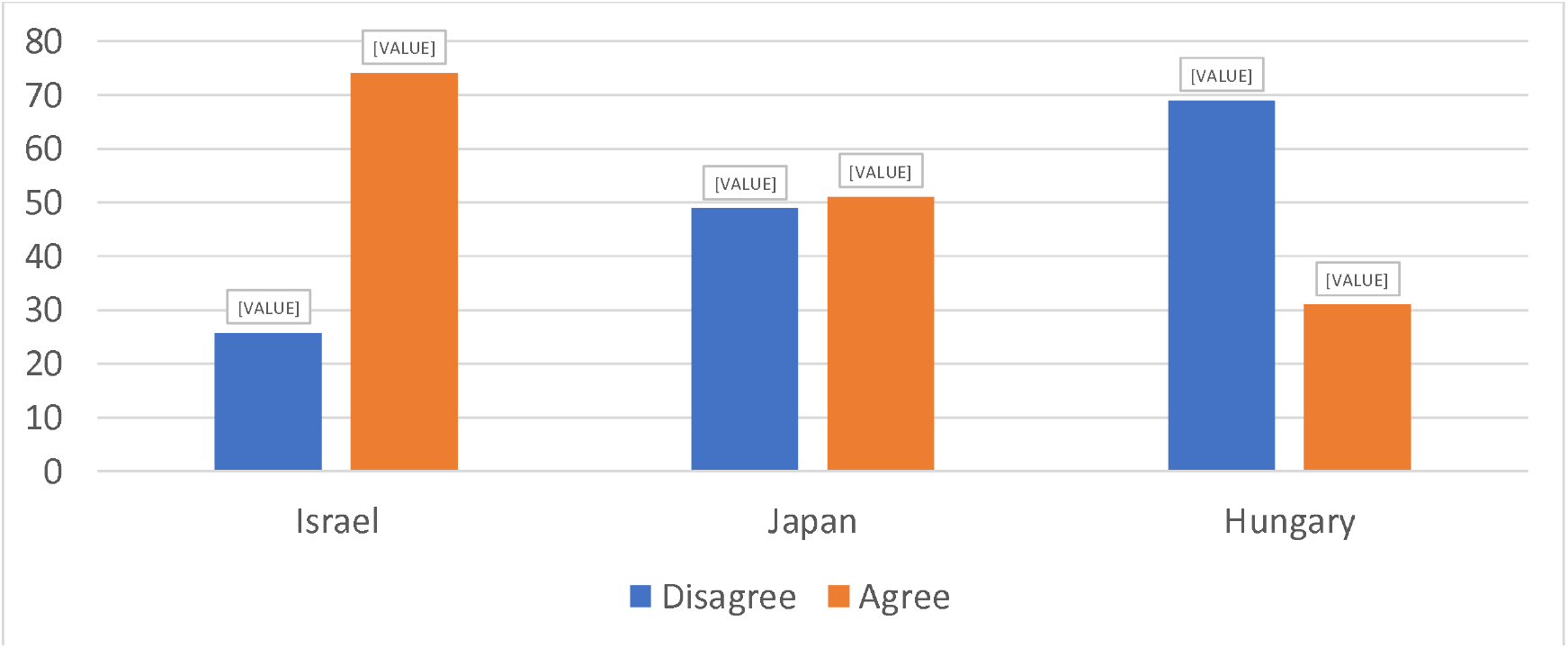
Proportion willing to accept COVID-19 vaccine.

### Objective 2: Willingness to vaccinate by demographics

In **Israel**, willingness to vaccinate was greater amongst those who were: (1) older (M ages 41.20 (agree) vs. 38.38 (disagree); t= -4.95 P<.001), (2) male (78% men vs. 71% women, χ2 (1) = 6.19, P=.01), (3) more educated (χ2 (2) = 12.90 P=.002) (4) employed (χ2 (2) = 6.03 P=.05) (employed were more likely to vaccinate (76%), compared to those were not employed (73%) or lost their job during the pandemic (66%)). There was no difference in willingness to vaccinate between those who belonged to a risk group (73.5% with a condition vs. 76.2% of those without (χ2 (1) = 0.70 P=.41) or between secular vs. non-secular respondents in vaccine willingness (χ2 (1) = 2.35 P=.13).

In **Japan**, willingness to vaccinate was greater amongst those who were: (1) older (M ages 47.23 (agree) vs. 44.23 (disagree); t= -3.23 .001), (2) male (57% men vs. 45% women, χ2 (1) = 13.20, p<.001), (3) more educated (57% University educated vs. 45% not University educated, χ2 (1) = 11.47 p=.001) (4) employed full time (χ2 (1) = 8.91 P=.003) (57% vs. those not employed full-time (47%)) (5) with underlying health conditions (62.6% with a condition vs. 48.3% of those without (χ2 (1) = 11.90 P=.001)

Finally, in **Hungary**, vaccine willingness was greater amongst those who were: (1) older (M = 52.72 (agree) vs. 49.49 (disagree); t= 3.16 P=.002) and (2) male (37.4% men vs. 25.6% women) χ2 (1) = 18.27, P=.001). Willingness was not significantly associated with education (χ2 (4) = 8.72 P=.068), but was associated with underlying health conditions (35.4% with a condition were willing to vaccinate vs. 28.1% of those without (χ2 (1) = 6.57 P=.01)).

### Objective 3: Testing an ecological model

We utilized layered multigroup logistic regression analysis using MPlus 8.1 to test our model of willingness to vaccinate. We included eight potential predictors of vaccine willingness: (i) likelihood of infection; (ii) Perceived severity of infection; (iii) Benefits of vaccination; (iv) Barriers to vaccination; (v) anticipated regret; (vi) Subjective norms (vii) trust in government. (viii) false vaccine beliefs. Age, gender, self-rated health (SRG), and risk group were included as covariates in the model, as were personal and family positive diagnoses of COVID-19.

In **Israel (**Table 2) results showed that, with the exception of sex (where women were less willing to vaccinate, adjusted odds ratio (aOR) = .58, P=.018) there was no significant association between demographic variables and vaccine intention (aORs = .97-.99). Higher subjective rated health was associated with reluctance to vaccinate (aOR = .66, P=.048). Participants who had been formally diagnosed with COVID-19 as well as family member who had been infected were not significantly associated with willingness to vaccinate. There were positive associations between willingness to vaccinate and the cognitive factors of *benefits of vaccine* (aOR = 2.11, P<.001), *anticipated regret* (aOR = 1.72, P < .001), while *barriers to vaccination* were associated with reluctance to vaccinate (aOR = .72 P < .001). *Perceived severity* and *perceived likelihood* were not associated with willingness to vaccinate, but *subjective norms* (aOR = 1.38, P = .008) was associated with more willingness to vaccinate. *Trust in government* was positively associated with willingness to vaccinate (aOR = 1.38, P = .002), *false beliefs about COVID-19* significantly associated with unwillingness to vaccinate (aOR = .11, P < .001).

**Table 2.**
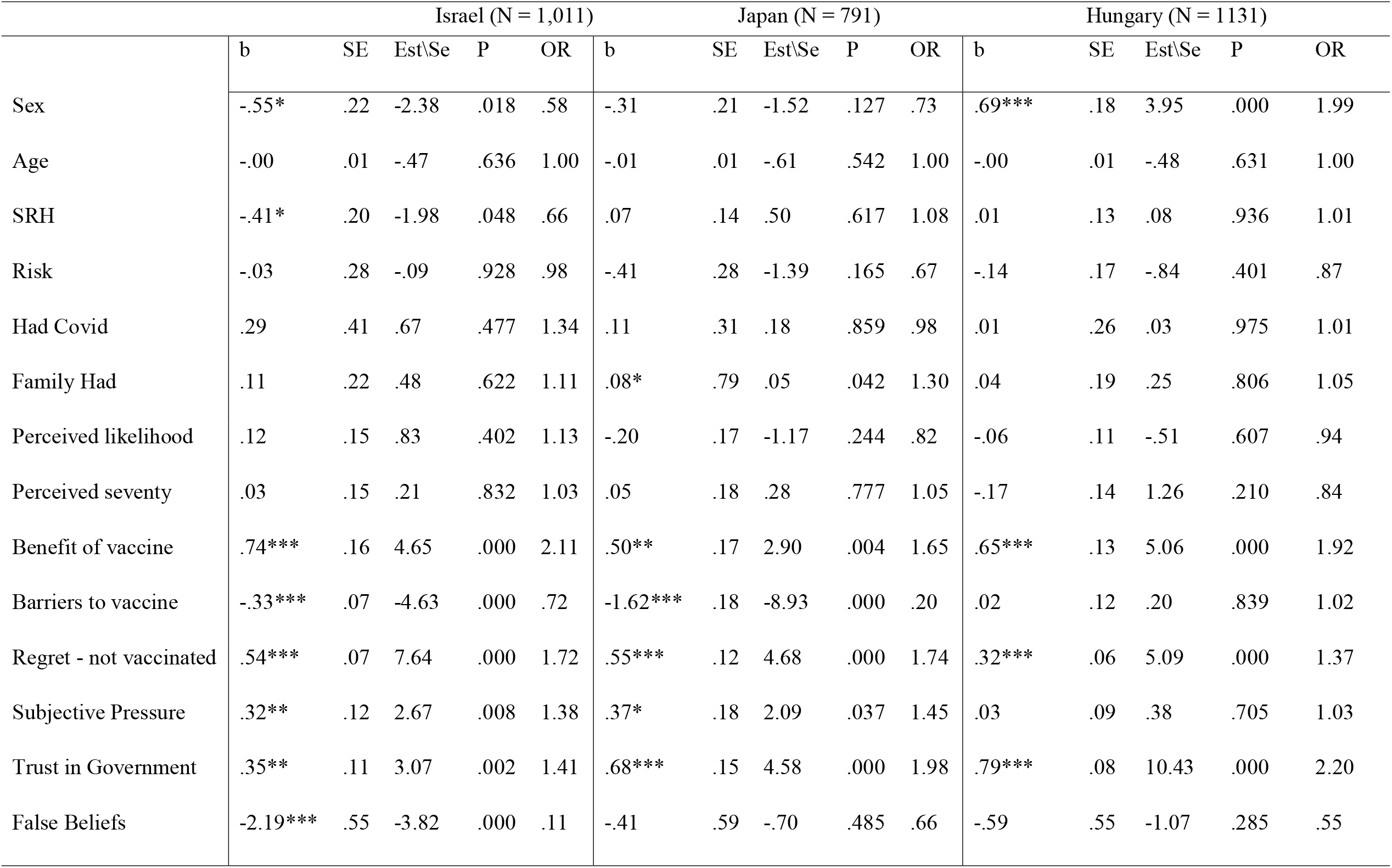
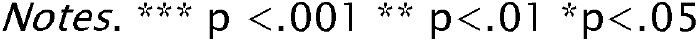
Magnitude, Statistical Significance and Odds-Ratio for Willingness to Vaccinate

|  | Israel (N = 1,011) |  |  |  |  | Japan (N = 791) |  |  |  |  | Hungary (N = 1131) |  |  |  |  |
| --- | --- | --- | --- | --- | --- | --- | --- | --- | --- | --- | --- | --- | --- | --- | --- |
|  | b | SE | Est\Se | P | OR | b | SE | Est\Se | P | OR | b | SE | Est\Se | P | OR |
| Sex | -.55* | .22 | -2.38 | .018 | .58 | -.31 | .21 | -1.52 | .127 | .73 | .69*** | .18 | 3.95 | .000 | 1.99 |
| Age | -.00 | .01 | -.47 | .636 | 1.00 | -.01 | .01 | -.61 | .542 | 1.00 | -.00 | .01 | -.48 | .631 | 1.00 |
| SRH | -.41* | .20 | -1.98 | .048 | .66 | .07 | .14 | .50 | .617 | 1.08 | .01 | .13 | .08 | .936 | 1.01 |
| Risk | -.03 | .28 | -.09 | .928 | .98 | -.41 | .28 | -1.39 | .165 | .67 | -.14 | .17 | -.84 | .401 | .87 |
| Had Covid | .29 | .41 | .67 | .477 | 1.34 | .11 | .31 | .18 | .859 | .98 | .01 | .26 | .03 | .975 | 1.01 |
| Family Had | .11 | .22 | .48 | .622 | 1.11 | .08* | .79 | .05 | .042 | 1.30 | .04 | .19 | .25 | .806 | 1.05 |
| Perceived likelihood | .12 | .15 | .83 | .402 | 1.13 | -.20 | .17 | -1.17 | .244 | .82 | -.06 | .11 | -.51 | .607 | .94 |
| Perceived severity | .03 | .15 | .21 | .832 | 1.03 | .05 | .18 | .28 | .777 | 1.05 | -.17 | .14 | 1.26 | .210 | .84 |
| Benefit of vaccine | .74*** | .16 | 4.65 | .000 | 2.11 | .50** | .17 | 2.90 | .004 | 1.65 | .65*** | .13 | 5.06 | .000 | 1.92 |
| Barriers to vaccine | -.33*** | .07 | -4.63 | .000 | .72 | -1.62*** | .18 | -8.93 | .000 | .20 | .02 | .12 | .20 | .839 | 1.02 |
| Regret - not vaccinated | .54*** | .07 | 7.64 | .000 | 1.72 | .55*** | .12 | 4.68 | .000 | 1.74 | .32*** | .06 | 5.09 | .000 | 1.37 |
| Subjective Pressure | .32** | .12 | 2.67 | .008 | 1.38 | .37* | .18 | 2.09 | .037 | 1.45 | .03 | .09 | .38 | .705 | 1.03 |
| Trust in Government | .35** | .11 | 3.07 | .002 | 1.41 | .68*** | .15 | 4.58 | .000 | 1.98 | .79*** | .08 | 10.43 | .000 | 2.20 |
| False Beliefs | -2.19*** | .55 | -3.82 | .000 | .11 | -.41 | .59 | -.70 | .485 | .66 | -.59 | .55 | -1.07 | .285 | .55 |

*Notes.* \*\*\* $p < .001$ \*\* $p < .01$ \* $p < .05$

In **Japan**, participants who had a family member infected and ill with COVID-19 were more willing to vaccinate (aOR = 1.30 p = .042), but neither participant’s own diagnosis with COVID-19 nor their demographic variables or health status was associated with vaccine intention. There were positive associations between willingness to vaccinate and the cognitive factors of the *benefits of vaccination* (aOR = 1.65, P = .004) and *anticipated regret* if not vaccinated (aOR = 1.74, P < .001) while *barriers to vaccination* were associated with reluctance to vaccinate (aOR = .20 P < .001). *Subjective norms* were associated with willingness to vaccinate (aOR = 1.45 p = .037). as was *trust in government* (aOR =1.98, P < .001), but not false beliefs (aOR=.66 P=.49).

Finally, in **Hungary**, findings indicated that, with the exception of sex (where men were more willing to vaccinate (aOR) = 1.99, P < .001)) there was no significant association between demographic or health status variables and vaccine intention (aORs = .99-1.00). Neither the participant’s nor their family’s formal diagnosis with COVID were related to willingness to vaccinate. There were positive associations between willingness to vaccinate and the cognitive factors of *benefits of vaccine* (aOR = 1.92, P < .001) and *anticipated regret* (aOR = 1.37, P < .001) but neither *barriers to vaccination, perceived severity or perceived likelihood of COVID-19*, nor *subjective norms* were associated with willingness to vaccinate. *Trust in government* was positively associated with willingness to vaccinate (aOR =2.20, P <.001) but false beliefs were not associated with such willingness.

### Objective 4: Clustering of beliefs across samples

A multi-group Latent Cluster Analysis tested one to four class solutions for the three samples, examining whether the solution demonstrated the same class pattern was obtained across samples^45^. As shown in Table 3, decrease in BIC was greatest for a three-class solution, providing very strong evidence of best fit [46](decrease > 10 times vs. a 5 time decrease for a subsequent class with no further theoretical justification for a fourth class). Relative entropy for a three-class model indicated good classification accuracy (.87 accuracy of class membership in any culture). In each country classes 1 and broadly 3 reflected either high or low false beliefs overall with an intermediate mixed belief category differing by country, with country classes most similar for Israel and Hungary. Class-specific conditional probabilities for each indicator are displayed in Fig. 3a-3c.

**Table 3:** Fit Indices for One-Four Multi-Group Latent Class Models

|  | **AIC** | **BIC** | **ssBIC** | **Entropy** | **(df) $\chi^2$** |
| --- | --- | --- | --- | --- | --- |
| **1-class** | 38632.94 | 38826.50 | 38724.83 | 1.00 | (2992) 17427.94 |
| **2-class** | 35454.51 | 35835.58 | 35635.40 | .91 | (3000) 6168.52 |
| **3-class** | **34925.15** | **35493.73** | **35195.06** | **.87** | **(2972) 4746.94** |
| **4-class** | 34679.88 | 35435.97 | 35038.80 | .85 | (2943) 3130.93 |

*Note.* Abbreviations: AIC = Akaike Information Criterion, BIC = Bayesian Information Criterion, ssBIC = Sample Size Adjusted, Bayesian Information Criterion

**Fig 3.**
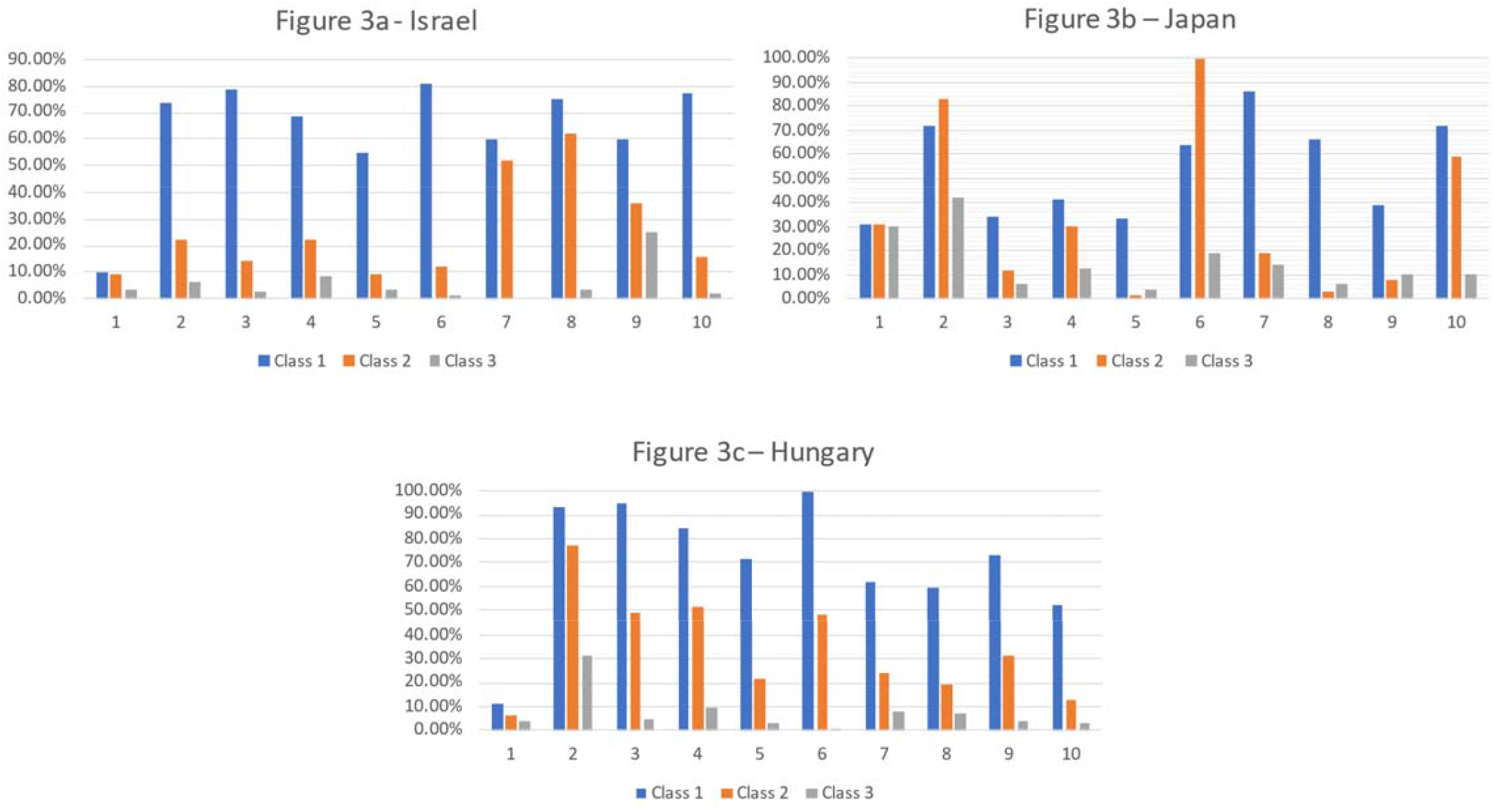
Three class solution of a Latent Profile Analysis of the False Belief scale. Note. Items - 1 - The flu vaccine will protect you against COVID-19; 2 - The COVID-19 vaccine causes allergies; 3 - Vaccines weaken the immune system (NHS); 4 - Vaccines do not cause autism (reverse coded); 5 - Vaccines do not contain mercury (reverse coded); 6 - The COVID-19 vaccine can give you covid- 19; 7 - If you have had COVID-19 already you can still benefit from the covid-19 vaccine (reverse coded); 8 - Receiving an mRNA vaccine will alter your DNA; 9 - The vaccine has severe side-effects; 10 - Reactions to the COVID vaccine are mild (reverse coded)

### Israel

Respondents assigned to Class 1 (**High False Beliefs**, n = 125, 12.3%) exhibited high probability for all items (60% - 81.30%) with the exception of the beliefs that *the flu vaccine protects against COVID-*19 (9.90%) and *the vaccine causes autism* (moderate probability, 54.60%). Those assigned to Class 2, a combination of **the vaccine may change DNA + overall Low False Beliefs** (n = 336, 33.2%) showed low probabilities for all false beliefs (8.90-36.30%) except for a medium-high probability that *the vaccine alters DNA* (62.30%) and that *people can(not) benefit from the vaccine even if they had COVID* (51.90%). The third class, **Low False Beliefs** (n = 550 54.4%) showed low probabilities for each belief (ranging from 0-25%).

Separate Logistic Regression showed Classes 1 and 2 were associated with greater unwillingness to vaccinate (vs. the low false belief reference group, class 3). (χ^2^(1) = 215.97 p < .001). **High False Beliefs** was most closely associated with unwillingness to vaccinate (b = -3.36 SE = .26 Wald = 170.86 p < .001 odds ratio = .035). **Vaccine may change DNA + Low False Beliefs** was also associated with unwillingness to vaccinate compared to the reference group (b = -1.14 SE = .18 Wald = 40.50, p < .001 odds ratio = .320).

### Hungary

Those assigned to Class 1 **High False Beliefs** (n = 136, 12.0%) exhibited high probability of all items (59.80% - 100%) except for the belief that *the flu vaccine protects against COVID-19* (low probability 10.80%) and that *reactions to the vaccine are mild* (moderate probability, 52.0%). Participants assigned to Class 2, **High belief in Allergies + overall moderate-Low False Beliefs** (n = 422, 37.3%) exhibited low probabilities for all false beliefs (6-31.50%) except for the belief that *the vaccine causes allergies* (77.40%) and the moderately-held beliefs *the vaccine weakens the immune system* (49.30%), *causes autism* (51.70%) and *can give you COVID-19* (48.00%). For the third class, **Low False Beliefs** (n = 573, 50.7%) probabilities of each belief were low (range .20-31%).

Separate Logistic Regression showed again that the first two classes were significantly associated with (un)willingness to vaccinate compared to the reference group (low false beliefs) (χ^2^(1) = 122.78 p < .001): for **High False Beliefs** (b = -2.84 SE = .43 Wald = 44.48 p < .001 odds ratio = .058); **High belief in Allergies + overall moderate-Low False Beliefs** (b = -1.04 SE = .14 Wald = 51.79, p < .001 odds ratio = .353).

### Japan

Respondents assigned to Class 1 - **High-Moderate False Beliefs** (n = 115, 11.6%) – were more likely to believe that *people can(not) benefit from the vaccine even if they had COVID-19* (86%), *the vaccine causes allergies* (72%), *the vaccine may give you COVID-19* (64%), *alter DNA* (66%), the vaccine has *mild side-effects* (72%). with other false beliefs at moderate levels (31.30 – 41.40%). Class 2, **Low beliefs + Vaccine can give you COVID-19** (n = 427, 43.2%) showed lower probabilities of all false beliefs (1.08-30.90%) with the exception of *the vaccine can give you COVID-19* (100%) or *the vaccine causes allergies* (83%) as well as moderate levels of belief that *reactions to the vaccine are mild* (59.40%). The third class Low False Beliefs (n = 446, 45.1%) had low probabilities for all beliefs (ranged 6.20-30.10%) except for the belief that *the vaccine can cause allergies* (41.90%).

Separate Logistic Regression showed that the neither Class 1 nor Class 2 of false beliefs were associated with willingness to vaccinate, when compared to the reference Class (3): (χ^2^(1) = 1.08 p = .297: class 1 (b = -.22 SE = .22 Wald = .99, p = 318 odds ratio = .800); Class 2 (b = -.09 SE = .14 Wald = .39, p = 531 odds ratio = .915).

## Discussion

Across the world there is evidence of continued vaccine unease, vaccine resistance identified as a top ten threat to global health even before COVID-19^47^. Countries however have fared differently in the availability of vaccines and the trust of their populations towards a growing range of possible vaccines, with a nested set of factors influencing uptake.

In Israel, a well-established community-based health service, strong logistic capacity, large-scale public health campaigns and the early purchase of a large number of vaccines helped the country achieve a rapid and comprehensive roll-out of the Pfizer-BioNtec vaccine^48, 49^. Unsurprisingly therefore almost three-quarters of our sample in this country demonstrated willingness to be vaccinated. In contrast, and despite greater personal exposure to the pandemic and perceived severity of threat, our Hungarian and Japanese respondents were less willing to vaccinate. In our sample only just over half (51%) of Japanese respondents indicated such willingness, higher than the 45% reported by the Imperial College COVID-19 tracker in January 2021^3^ but lower than the 62% indicated by a cross-sectional survey also conducted that month^17^. In Hungary, the politicisation of the vaccine roll-out^7^, and disputes over the use of vaccines not approved by the EU^12^, contributed to high national rates of scepticism about efficacy^50^ with only 31% of our national sample expressing willingness to vaccinate, and a further 21% uncertain.

Across our three country samples willingness to vaccinate was greater amongst older respondents, the age group most at risk of mortality/ morbidity from the SARS-CoV-2 virus, consonant with previous research ^32, 36^. In further bivariate analyses in Japan and Israel the more educated and the employed were also more likely to vaccinate^36^. Rejection of covid-19 myths was associated with increased years schooling in Hungary (r = -.24 P=.001) or being a graduate in Israel (t (722) = -4.79 P=.001), suggesting that the more educated challenge some of the myths that contribute to vaccine hesitancy or rejection. This is consistent with cross-cultural evidence suggesting the more numerate are less suspect to COVID-19 misinformation^32^. In Japan and Hungary those with an underlying health risk were more willing to vaccinate, in line with previous surveys on influenza vaccination^50^. In competitive regression which included all three levels of factors in our ecological model, however, demographic factors, individual disease vulnerability, and personal or family experience with COVID-19 had only a small association with vaccine willingness. Instead, in each culture, those who appreciated the benefits of the vaccine, those more likely to regret vaccinating, and respondents who trusted their governments and health authorities were more likely to indicate vaccine willingness. There were, however, only small and culturally variable associations between perceived likelihood or severity of infection and vaccine intention, indicating only a weak association between perceived threat and vaccination. This may be because while infection likelihood and severity are closely associated with viral threat, benefits and regrets may be more proximally associated with actual vaccine behaviour. Subjective pressure to vaccinate was significantly higher in Israel and Japan compared to Hungary and associated with vaccine uptake in just these first two countries. This suggests that the importance of friends, families and others for vaccine willingness may be particularly significant where important others are prepared to be vaccinated.

*Engagement and communication* were further, additional predictors of vaccine willingness. False information about the virus, most likely to emerge from social media, has been shown to be negatively associated with COVID-19 health protective behaviour^38^, including vaccine willingness^20^. In bivariate analyses in each country, false beliefs were negatively associated with vaccine uptake (rs = -.53 (Israel), -.29 (Japan), -.32 (Hungary), all P< .001). While these false beliefs varied significantly by group, in each country they were negatively related to age (rs -.12 (Israel), -.17 (Japan), -.13 (Hungary) P=.001 in each country). In Israel, where we included religion, false beliefs were also strongest in the Ultra-Orthodox population, lowest in secular respondents (F (3, 1003) = 4.68 P=.003). As reported elsewhere^31, 43^ these misbeliefs were significantly associated with low trust in governmental authorities (rs -.25,-.24 and -.32 for Israel, Japan and Hungary respectively, P=.001) but only the association in Israel between false beliefs and vaccine willingness survived the competitive regression models. Trust in government emerged in all three countries as a significant contributor to vaccine willingness, as elsewhere^36^. This association was strongest in Hungary, where vaccine uptake, and choice of vaccine, has been particularly politically contentious. Polling data collected co-temporaneous to our Hungarian study showed political divisions between those willing to vaccinate with a Hungarian government approved vaccine (including Russian and Chinese vaccines at that time not approved by the European Medicines Agency(EMA)) and those on the political left more likely to endorse the EMA recommended Pfizer or Moderna vaccines^51^.

In our samples, misbeliefs about the vaccine correlated with each other, supporting the idea of a ‘monological belief system’^32, 52^. Nevertheless, our latent class analysis allowed us to classify the structure of beliefs in each culture and their association with vaccine willingness, permitting a more nuanced understanding of how different groups may understand potential vaccine risks. In Israel and Hungary beliefs were similarly structured in classes, with one group of respondents (approximately 12%) demonstrating high rates of false beliefs and another (approximately half the sample) scoring low on false beliefs. In both countries high levels of false belief were associated with unwillingness to vaccinate, in accord with our regression analyses. However, in Israel the belief that *COVID-19 can alter your DNA* (held by 62% of those in the second category) was distinctive as a predictor of vaccine (un)willingness. Contrastingly, in Hungary it was the association between allergies and the vaccine (held by 77% of those in class 2) that distinguished a grouping of respondents reluctant to take the vaccine. In Japan, classes identified were generally more blended and less distinguishable, and subsequently less clearly relate to vaccine willingness. Despite this, all members of the largest latent class in Japan (43% of respondents) indicated their belief that *the vaccine can give you COVID-19*. Notably, 61.5% of Japanese respondents who believed *the vaccine can give you COVID* in Japan were unwilling to vaccinate, compared to 48.6% Israel and 14.0% in Hungary, suggesting the particular significance of this misbelief in Japan for vaccine intentions.

### Limitations

Our studies benefitted by including national samples from three very different cultures, at different stages in their vaccination programmes and with different histories of vaccine uptake. We go beyond most literature on vaccine uptake and hesitancy by examining vaccine willingness in an expanded model incorporating different ‘levels’ of influences: demographic, cognitive and societal, and by examining in more detail the structure of misbeliefs about the vaccine, by culture. However, we recognise a number of limitations to our survey. First, samples were cross-sectional, and were therefore not able to assess predictors of vaccine willingness over time. Data was first collected in early January, at the start of the first major vaccine roll-out, meaning that we did not include later misbeliefs that emerged in subsequent months and which often focused on the association between vaccination and government/’big Tec’ control and monitoring. This may be particularly importance with the arrival of new variants of concern that have challenged potential vaccine efficacy. In addition, emergent concerns over vaccine safety (such as worries about blood clotting following the AstraZenica vaccine^53^) may serve to directly inhibit uptake and perpetuate further new misbeliefs and distrust. Second, because of the speed of the evolving vaccination situation in both countries (the rapid programme vaccination in Israel, the introduction of vaccination in Japan) our survey companies expedited data collection within a short time period. Although widely used, and important particularly for the collection of time-sensitive data during a vaccination campaign, we recognise that the quota sampling we employed has important limitations in ascertaining accurate response rates^20, 54^. We distributed a large number of invitations at the potential expense of response rate and could not readily estimate non-response rate from those who saw the survey invitation. Our samples, while representative of the demographic characteristics of their wider populations, were not genuine probability samples, and particular caution is needed when making cross-cultural comparisons. In Hungary in particular the percentage of willingness to vaccine may have been underestimated by the omission of those already vaccinated. Third, we did not obtain information on occupation, despite significant variations by profession in exposure to COVID-19, which may have further impacted on vaccine willingness^55^, and we did not assess income, although those with lower income are less likely to indicate vaccine willingness^17^. Research in the UK suggest there is also likely to be variation by ethnicity in the misbeliefs that might mitigate against vaccination^44^. Fourth, we assessed only three countries, and future work should expand the testing of nested models across settings. Such expanded analyses can better assess the cultural values that play a further part in health protective behaviours during COVID-19^56^, with individualism helping encourage conspiracy beliefs^57^. Finally, we measure only behavioural intentions rather than actual vaccination behaviour. Although the link between the two has been well established^58^ we recognise that attitudes towards any vaccination are likely to vary as populations acquire further experience with the virus and the vaccination rollout. Future longitudinal research could profitably examine the impact of cognitive and broader cultural factors on consistency of vaccine commitment and resilience to new counter-messages, as well as behaviours post-vaccination, particularly in those critical weeks before full immunity is attained.

### Implications for vaccine drives

Despite these limitations we believe our findings have important implications as other nations strive to accelerate their vaccination drives. Encouraging vaccine uptake requires a planned approach involving consistent, credible communications that emphasise vaccine benefits, confronts potential barriers, and warns of potential later regret if the vaccine is not accepted. Vaccine campaigns may need not focus on disease threat: instead, these initiatives would better focus on the effectiveness of the vaccine, confront misinformation, and seek to emphasise the trustworthiness of key actors, such as national health services. Those vaccinated should be encouraged to inform close others, in order to emphasise the normative nature of this activity. Public health agencies need to reach people beyond remote media campaigns and be present where individuals shop and work^59^; doctors have been widely reported to be important in addressing myths^39^ with pharmacists in Hungary significant for encouraging influenza vaccination in that country^50^. Social media is of course likely to have an important role, with vaccine resistance highest amongst who obtain their information from such media^20^, although source of the media is important (e.g. obtaining information from the WHO was associated with lower susceptibility to COVID-19 myths^32^; those who use Instagram or What’sApp for information on the pandemic are more likely to believe that the vaccine negatively impacts the immune system than those using traditional media^44^). Our analyses suggested that, despite some similarities in belief structure, there were distinctive beliefs in each culture important for understanding vaccine willingness. In Israel concerns need to be addressed about the possibility of DNA being altered by the (mRNA) vaccine employed; for Hungary vaccine campaigns need to be particularly wary of concern about allergies while in Japan such interventions need to be alerted to the belief that *the vaccine can give you COVID-19*. Sensitivity to these particular beliefs in one culture may be particularly important where views on vaccination reflect political divisions: our Israeli data was collected shortly before a highly contested election where vaccination roll-out was a major campaign topic, in Hungary the use of vaccines yet to be approved by European Medicines Agency caused controversy^60^, while the Japanese data was gathered only a few months before a controversial Olympic Games^61^, with vaccination a national priority in the run-up to this event. Finally, within country, group factors may be also particularly important. Younger respondents need to be encouraged in particular to vaccinate; this may require addressing in particular COVID-19 myths most prevalent in the social media consumed by younger audiences^40^. Two weeks after our data collection (1st Feb 2021), actual vaccination uptake in Israel amongst those aged over 60 varied significantly, with 66.1% uptake in the ultra-Orthodox community and 60% in the Arab population, compared to 84.9% in the wider populace^20, 61^. Uptake was also greater in settlements with higher socio-economic status, despite the greater morbidity from COVID-19 amongst poorer communities^62^. To address these variations in uptake the specific concerns of religious and other social groups need to be considered, with community leaders actively engaged through culturally appropriate conversations in order to allay fears, address specific myths and thus help further facilitate a successful vaccine campaign^63^.

## Methods

### Participants and procedure

Data were from nationally representative samples of adult populations collected in Israel (N = 1011), Japan (N = 997) and Hungary (N=1131), using large panel survey companies in each country (iPanel for Israel, Quesant! for Japan, Medián Opinion and Market Research in Hungary). In each case quota sampling was used to ensure the sample: participant sex, age (Israel), sex, age and geographical region (Japan, Hungary) were chosen to match population parameters in each country. Data in Israel were collected from 31.12.2020 to 11.01.2021 i.e. early on during the first vaccination drive. At that time the percentage of those receiving their first vaccination doubled, from 11% to 22% of the population^2^. In Hungary, data were obtained from 8.4.21 to 16.4.21, during which 27.8% (then 32.0%) had received at least one vaccine^2^. In Japan data were collected prior to the start of the vaccination campaign (between 15.2.2021 and 16.2.2021).

Because we were keen to obtain samples either early in vaccination campaigns (Israel, Hungary) or prior to its commencement (Japan) time sensitivity meant requesting the survey companies to obtain samples of 1000 respondents in each country. This resulted in sample sizes comparable to those gathered in several major international studies of vaccine willingness^3, 64^. For each sample participants were contacted as part of cloud panels administered by the survey company and asked to participate via email. They were then reimbursed by the companies for their participation. In each country inclusion criteria required participants be the approved minimal age set by ethical requirements (Israel, Hungary - 18, Japan - 20), and to successfully pass validation checks (both specific items and timing of responses) to ensure participant attention and accuracy. All respondents were fluent in the relevant national language (Hebrew, Japanese, Hungarian). Ethical approval was from Ariel University’s Institutional Review Board (No. AU-SOC-MBE-20201224), the Yamaguchi University Review Committee for Non-Medical Research Involving Human Participants (2020-004-01) and the Eötvös Loránd University ethical review panel (PPK KEB 2021/130-2).

## Measures

### Demographics

Participants in each country indicated their age and sex. Because demographic information procedures vary across countries we obtained slight variations in each country, in common with other such cross-cultural comparisons^20^, while retaining the core ecosystems model variables in each country for comparative analysis. In Japan and Israel respondents identified whether they completed only high school prior to University or were currently a student/had graduated; in Hungary students indicated their level of schooling (up to secondary school (N=44 (3.9%); secondary school (N=647 (57.3%), college degree (N=276, 24.4%), masters’ degree (N=147, 13.0%) or doctorate (N=16, 1.4%). In Israel respondents indicated if were employed (71.7%), unemployed (15.8%) or lost employment due to the pandemic (12.5%); in Japan only whether or not they were employed. Table 1 provides further descriptive statistics for each country.

### Health conditions

Respondents indicated their risk group membership using the US CDC risk group memberships (e.g. hypertension, diabetes). Participants also indicated whether they had been formally diagnosed with COVID-19 (yes, no), whether someone from their social circle had been thus diagnosed (yes, no), and their self-rated health (from bad (1) to excellent (4)).

### Vaccine acceptance

Participants were asked *Would you be willing to accept a vaccine approved safe and effective by the government?* (strongly disagree to strongly agree)). Because we questioned respondents at the start of actual vaccine campaigns, rather than about a hypothetical vaccine, and were cautious about both the translation of ‘uncertain/undecided’ and the use of intermediate responses in some cultural contexts^65^) we treat this as a binary response, in line with much previous cross-national vaccine research^36^. Responses were classified as 1 (‘completely agree’ or ‘somewhat agree’) vs. 0 (undecided, unwilling, very unwilling)^3, 36^.

### Predictors of vaccine willingness

We included eight potential predictors of vaccine willingness drawn from three major theoretical perspectives previously used to assess vaccine uptake: the Health Belief Model (HBM), the Theory of Planned Behaviour (TPB) and the Theory of Protection Motivation (TPM)^18, 19, 58^, as well as subsequent work on the influence of false beliefs and trust in authorities on vaccine willingness. Full items and response categories are reported in the Online Appendix, scale alphas in Table 1.

The first six sets of questions were drawn from the HBM and TPM and employed questions previously used to assess swine flu vaccination^18, 19^.

*Likelihood of infection*. Here we asked three questions on likelihood of infection *e*.*g. Getting COVID-19 is currently a possibility for me*, which previously showed acceptable scale reliability for assessing vaccination intentions^19^ *Perceived severity of infection*. We use a further 3 items drawing on the HBM and TPM e.g. *I will be very sick if I get COVID-19*. These also showed acceptable scale reliability for assessing vaccination intentions^19^

*Perceived benefits of vaccination*. These three items included *Vaccination is a good idea because I feel will be less worried about catching COVID-19*^19^

*Barriers to vaccination* indicate two daily impediments to vaccination e.g. *The side-effects of COVID-19 will interfere with my usual activities*^19^

*Anticipated regret* is a single item drawn from an extension of TPB^19^ and was previously a significant predictor of intention to be vaccinated for 2009 H1N1*(If I did not have a COVID vaccination, I would later wish I had)*

*Subjective norms* used items taken from the TPB with previous good reliability^19^. These indicate influence of others on willingness to be vaccinated, and was assessed by five items (e.g. *My friends would approve of me having the COVID-19 vaccination*)

*Trust in government* extended a similar item used in a previous global study of vaccine acceptance during COVID-19^36^ and included three items to assessing generalised trust, trust related to COVID-19 and trust with respect to vaccination (*e*.*g. I trust Government regarding vaccination*)

*False beliefs* were a set of ten true or false items taken from the myth-busting websites of the WHO, CDC and the UK NHS. Four of these were reverse coded. These included *The COVID-19 vaccine can give you covid-19* (from the NHS website), and *receiving an mRNA vaccine will alter your DNA* (CDC)

### Analytic procedure

We utilized layered multigroup logistic regression analysis using MPlus 8.1^66^ to test the ecological model. Data were analysed using maximum likelihood estimation with robust standard errors (MLR) to handle non-normal distributions. We used multigroup analyses^67^ to test if paths from the covariates, demographic factors and health status, individual cognitions, normative pressures, trust in government, belief in COVID-19 myths and willingness to be vaccinated varied by culture. We included participant information on their own (or social network’s) positive COVID-19 diagnoses. Missing data due to nonresponse was minimal, ranging from .9% to 4%.

To analyse the structure of misbeliefs about the vaccine we employed Multi-Group Latent Class Analysis (MLCA) to identify subgroups within the three samples (Israel, Japan and Hungary). We use binary indicators of false beliefs. We specified models with one to four classes and compared the models to determine the optimal number of classes. Models with lower values on the Akaike Information Criterion (AIC), Bayesian Information Criterion (BIC), and sample-size adjusted Bayesian Information Criterion (aBIC) were prioritized. Entropy values and average latent class probabilities indicated classification accuracy, with preference for models with entropy values and probabilities of correct class assignment closer to 1.00^68^. Once optimal number of classes was determined we computed sample percentages assigned to each class and conditional probabilities by class, with labels for latent classes based on patterns of probabilities for endorsing each false belief.

## Data Availability

The datasets generated and/or analysed during the current study are available in the Open Science Framework (OSF) repository, and can be accessed here: https://osf.io/dm587/.

## Acknowledgments

Funding for the study in Israel was awarded to Prof. Ben-Ezra from Ariel University No. RA2000000522 in Japan to Professor Masahito Takahashi (JSPS KAKENHI Grant Number 18K01963) and in Hungary to Drs. Lan Anh Nguyen Lu and Mónika Kovács (National Excellence Programme 2018-1.2.1-NKP-2018-00006).

## Author contributions

Conceptualization: MBE, RG, WHK, MT, LNL Methodology: RG, MT, MBE, YL, WHK, LNG Formal analysis and investigation: YL, RG, MBE, KB, MK, YHR, KB, MK Writing - original draft preparation: RG, YL, MBE, WHK; Writing - review and editing: all authors, Funding acquisition: MBE, MT, LNL, KB, MK; Supervision: RG, MT, MBE, YHR, LNL.

## Supplementary material

Items included in the questionnaire.

Scale, Response options

## Likelihood of infection ((1) not at all to (5) very much)

My chance of getting COVID-19 in the next few months is great

I am worried about the likelihood of getting COVID-19 in the near future

Getting COVID-19 is currently a possibility for me

## Perceived severity of COVID-19 infection ((1) not at all to (5) very much)

Complications from COVID-19 are serious,”

I will be very sick if I get COVID-19

I am afraid of getting COVID-19

Benefits of vaccine ((1) not at all to (5) very much)

Vaccination is a good idea because I feel will be less worried about catching COVID-19

Vaccination decreases my chance of getting COVID-19 or its complications

If I get vaccinated I will decrease the risk of spreading the disease to others

## Barriers to vaccine ((1) not at all to (5) very much)

The side-effects of COVID-19 will interfere with my usual activities

I cannot be bothered to get a COVID flu vaccination

## Anticipated regret ((1) strongly disagree to (7) strongly agree)

If I did not have a COVID vaccination, I would later wish I had

## Subjective norms ((1) strongly disagree to (7) strongly agree)

People who are important to me would approve of me having the COVID-19 vaccination

My family would approve of me having the COVID-19 vaccination

My friends would approve of me having the COVID-19 vaccination

I feel under pressure to have a COVID-19 vaccination

People who are important to me influence my decision to have the COVID-19 vaccination

## Beliefs (1(no) 2 (yes) (R) indicates reverse coding

The flu vaccine will protect you against COVID-19

The COVID-19 vaccine causes allergies

Vaccines weaken the immune system

Vaccines do not cause autism (R)

Vaccines do not contain mercury (R)

The COVID-19 vaccine can give you covid-19

If you have had COVID-19 already you can still benefit from the covid-19 vaccine (R)

Receiving an mRNA vaccine will alter your DNA

The vaccine has severe side effects

Reactions to the COVID vaccine are mild (R)

## Trust in government (1) not at all to (5) very much)

I trust the Government in general

I trust Government to deal with COVID-19

I trust Government regarding vaccination

